# COVID-19 AND PATIENTS UNDERGOING PHARMACOLOGICAL TREATMENTS FOR IMMUNE-MEDIATED INFLAMMATORY DISEASES: PROTOCOL FOR A RAPID LIVING SYSTEMATIC REVIEW

**DOI:** 10.1101/2020.05.01.20087494

**Authors:** Aline Pereira da Rocha, Alvaro Nagib Atallah, Ana Carolina Pereira Nunes Pinto, César Ramos Rocha-Filho, Felipe Sebastião de Assis Reis, Keilla Martins Milby, Vinicius Tassoni Civile, Nelson Carvas Junior, Rodolfo Rodrigo Pereira Santos, Laura Jantsch Ferla, Giulia Fernandes Moça Trevisani, Gabriel Sodré Ramalho, Maria Eduarda Santos Puga, Virgínia Fernandes Moça Trevisani

## Abstract

**CONTEXT AND OBJECTIVE:** We propose to systematically review the available evidence to evaluate if patients with immune mediated inflammatory diseases under pharmacological treatment with immunosuppressants, immunobiologics, Disease-Modifying Anti-Rheumatic Drugs (DMARD) or targeted synthetic DMARDs have better or worse outcomes when infected by severe acute respiratory syndrome coronavirus-2 (SARS-CoV-2). This study is a protocol for our rapid living systematic review. METHODS: Protocol for a rapid living systematic review methodology following the Preferred Reporting Items for Systematic Review and Meta-Analysis Protocols (PRISMA-P) guidance. To conduct the rapid systematic review, we will employ abbreviated systematic review methods, including: not performing independent screens of abstracts and not searching grey literature. As this will be a living review, it will be continuously updated.

## INTRODUCTION

These days, the world is facing the severe acute respiratory syndrome coronavirus-2 (SARS-CoV-2) emergency, characterized in March by the World Health Organization as a pandemic (1). It is well established in the literature that patients with compromised immune systems are more susceptible to viral infections and consequent development of severe outcomes compared with the general population. Patients with rheumatic diseases, for example, are known to present an increased infectious risk due to the disease itself and also to the iatrogenic effect of immunosuppressive agents (3). In addition, the presence of comorbidities often observed in these patients contributes to poor outcomes in viral infections (2).

In contrast to this evidence, some relevant clinical reports have described patients SARS-CoV-2 positive undergoing immunosuppressed treatment with mild symptoms and/or no severe outcomes (3). In fact, the reports have discussed that immunosuppression may protect COVID-19 patients from clinical complications.

Based on the knowledge of other viral infections, physicians and researchers hypothesize that antimalarial treatments, such as chloroquine, may prevent SARS-CoV-2 infection through endocytosis (4). Moreover, it is suggested that conventional and biological Disease-Modifying Anti-Rheumatic Drugs (DMARDs) may control pro-inflammatory cytokine expression and limit tissue damage (4). Despite the propositions, many aspects of these therapies in COVID-19 continue to be pending matters.

To address these gaps, we propose to systematically review the available evidence to answer the following question: “Do patients with immune mediated or inflammatory diseases under pharmacological treatment with immunosuppressants, immunobiologics, DMARDs or targeted synthetic DMARDs have better or worse outcomes when infected by SARS-CoV-2?” This study is a protocol for our rapid living systematic review.

## METHODS

This rapid living systematic review protocol will be developed following the Preferred Reporting Items for Systematic Review and Meta-Analysis Protocols (PRISMA-P) guidance (5), and was registered in the PROSPERO “International Prospective Register of Systematic Reviews” (CRD42020179863). To conduct the rapid systematic review, we will employ abbreviated systematic review methods, including: not performing independent screens of abstracts and not searching grey literature (6). As this is a living review, it will be continuously updated.

### Design

We will perform a rapid living systematic review methodology following the recommendations proposed by the Cochrane Collaboration Handbook (7).

### Eligibility criteria

#### Types of studies

We will include randomized controlled trials (RCT), quasi-RCTs, cohort, case-control studies, case series, and electronic health records that evaluated the effects of immunomodulatory drugs in patients with immune-mediated inflammatory diseases infected by SARS-CoV-2.

#### Types of participants

We will include studies with patients with confirmed diagnosis of infection of SARS-CoV-2 and immune-mediated inflammatory diseases.

#### Type of interventions

- Immunosuppressants (e. g. methotrexate, azathioprine, mycophenolate; cyclophosphamide);
- Immunomodulators (e.g glucocorticoids; immunoglobulins);
- Immunobiologics (e.g tocilizumab, infliximab, adalimumab, etanercept, certulizumab, rituximab, secukinumab, ustekinumab);
- DMARDs (e.g chloroquine, hydroxychloroquine, sulfasalazine);
- Targeted synthetic disease-modifying anti-rheumatic drugs (e.g. apremilast, tofacitinib, baricitinib).

#### Outcome measures

- Primary outcomes
  - Mortality rate;
  - Length of hospital stay;
  - Adverse events;
- Secondary outcomes
  - Duration of invasive mechanical ventilation;
  - Time to viral clearance;
  - Time to clinical improvement;
  - Length of Intensive Care Unit stay;

#### Report characteristics

We will include studies performed since November 2019. No language restrictions will be used in the selection.

### Data sources and searches

We will search PubMed, Embase, Cochrane Central Register of Controlled Trials (CENTRAL), LILACS, Scopus and SciELO using relevant descriptors and synonyms, adapting the search to the specifications of each database to identify published, ongoing, and unpublished studies. We will also search the following COVID-19 specific databases: Epistemonikos COVID-19 L·OVE platform; ClinicalTrials.gov; The World Health Organization International Clinical Trials Registry Platform (WHO ICTRP). Finally, we will search the lists of references of the included studies. No language restrictions will be used in the selection.

### Search strategy

We will use the terms related to the problem of interest, the intervention and the filter the date of publication. The search strategy in Medline via Pubmed is shown in Table 1.

**Table 1.**
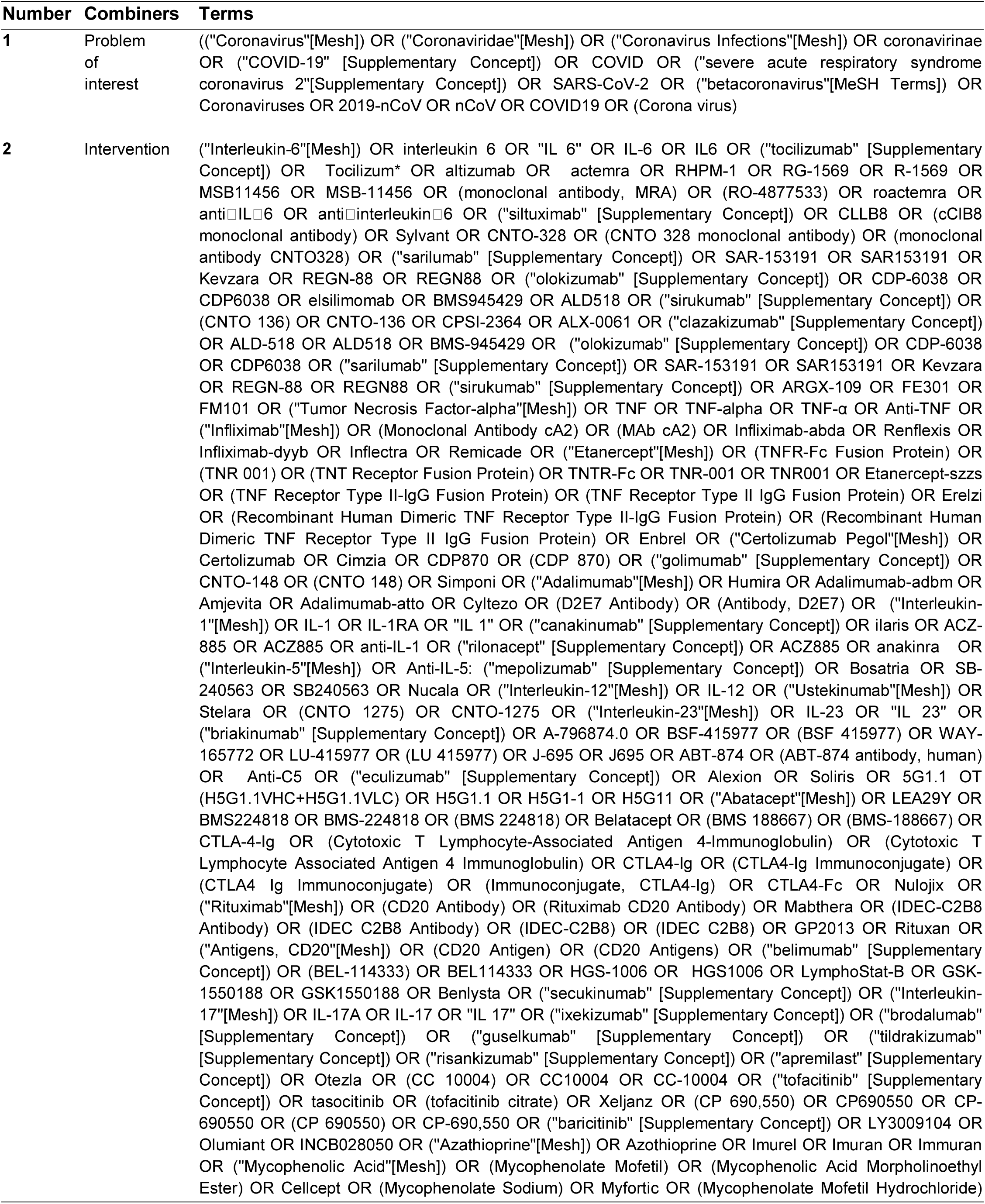
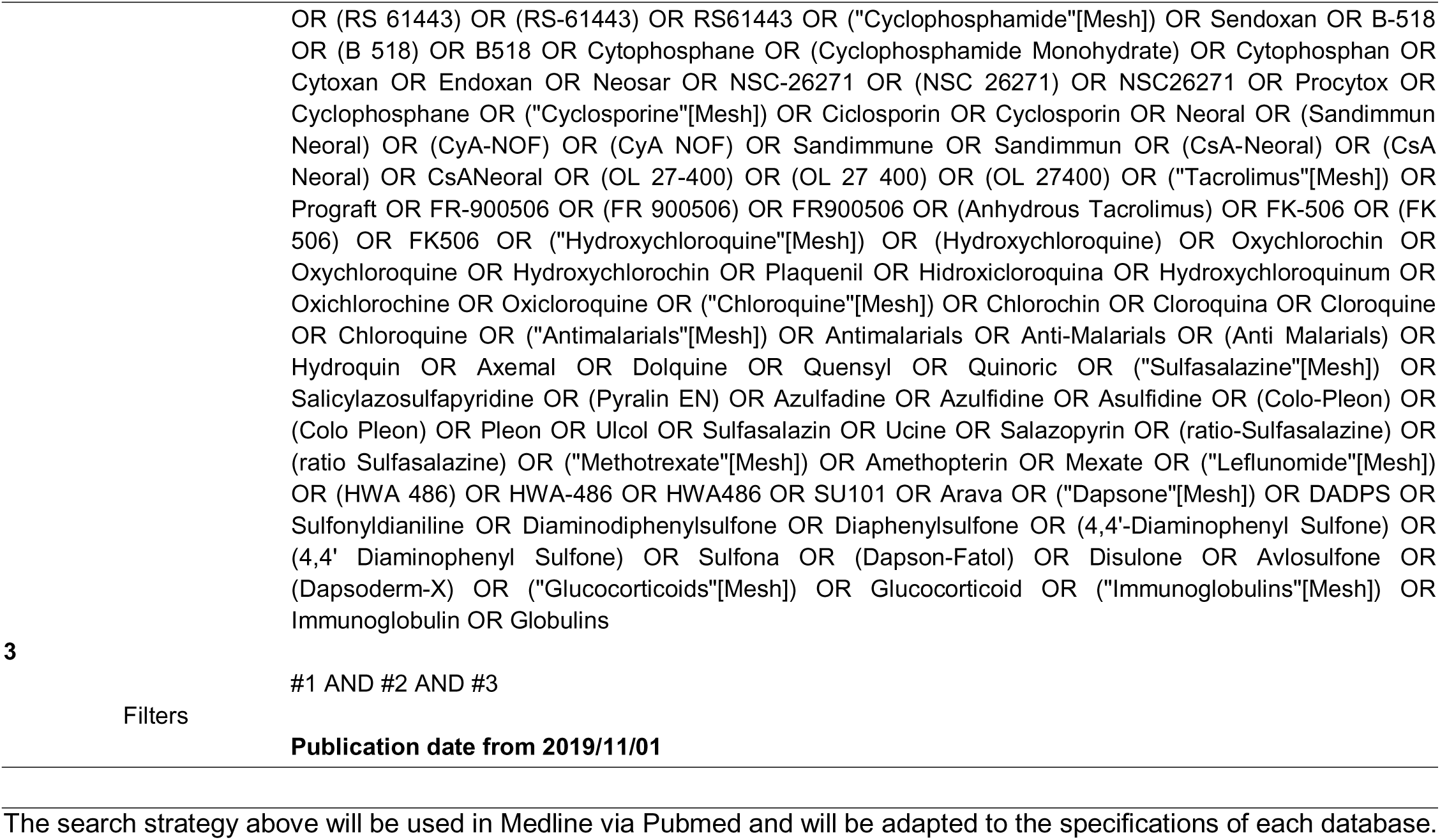
Systematic review Search Strategy

### Study selection

Two authors will select the studies for inclusion in the review (ACPNP and FSAR). When duplicated studies are found, only one of them will be considered for inclusion. When reports using the same participants and different follow-up or outcome measurements are found, both reports will be included. However, they will be considered as parts of only one study. After removing duplicated studies and/or reports, the authors will read the study titles and abstracts. Studies that clearly do not fulfill the eligibility criteria will be excluded. The remaining studies will then be fully read and assessed for inclusion in the review. Disagreements between authors regarding the inclusion of studies will be solved by the third author (VFMT). We will present the reasons for exclusion of the studies in a flowchart. To optimize the process of screening and selection of studies, Rayyan application (8) will be used.

### Data extraction

Two authors (APR and FSAR) will independently extract data. Discrepancies or disagreements will be solved by a third author (VFMT). We will use a predefined form to extract data from included studies. The form will include information on: - the patients (demographic and clinical characteristics); - time points used for the assessments; - the pharmacological treatment (name of drug, treatment duration; dose); - number of patients lost to follow-up (in each group); - reasons for loss to follow-up; - approach for handling missing data (data imputation/how data imputation was performed, use of intention-to-treat approach); - sources of funding; - possibility of conflict of interests; - adverse events; - outcome measures; - protocol deviations.

### Assessment of methodological quality in included studies and quality of the body of evidence

We will use Risk of Bias tool (7) as recommended by Cochrane Collaboration to perform the critical appraisal of included studies; for observational studies, we will use Newcastle Ottawa Scale (9). The quality of evidence will be assessed using the Grading of Recommendations Assessment, Development and Evaluation (GRADE) (10). Assessment of risk of bias (VTC and APR), and assessment of the quality of evidence (VTC and NCJ) will be performed by two authors, and all the disagreements in the assessment of the risk of bias or quality of evidence will be solved through discussion or, if required, by consulting with a third author (ANA).

### Data Analysis

We will perform analyses according to the recommendations of Cochrane, and the Cochrane Prognosis Methods Group. To perform the meta-analysis we will use R software when possible for mean difference (MD) or Hedge’s/Cohen’s (SMD). The package to be used is the “meta” (version 4.11-0).

We will pool mean and the standard deviation (or equivalent) for hospital admission, intensive care unit admission and/or respiratory support for adult inpatients with COVID-19 and mortality, by the inverse of variance method with random-effects model (DerSimonian-Laird estimator for τ2). Thus, just similar or equivalent results obtained by each primary outcome in the studies can be used. It is necessary to ensure the main assumptions to provide the correct data and to guarantee the correct interpretation. When allowed by the information of the studies results the correct statistical standardization will be provided. Aftermath, just the robust and reliable data based on equivalents primary outcomes can be used to attend the objective of this meta-analysis.

All described in the Data Extraction section for each measure must be separated according to the response categories of the interest. The data about the adverse events and clinical exams can be extracted since complete available and classified by the response as well.

### Dealing with missing data

For studies that do not provide a mean and associated standard deviation (SD), we will use information and results reported in the text or tables, doing the correct inference. When the parameters established before are not available, the estimate based on other parameters will be made ensuring the correct information.

We will contact the principal investigators of the included studies asking for additional data or to clarify issues about the studies. In the absence of a reply from the authors, we will expose the data in a descriptive manner avoiding imputation.

### Assessment of heterogeneity

We will employ the Cochran’s Q test to assess the presence of heterogeneity considering a threshold of P value < 0.1 as an indicator of whether heterogeneity is present.

In addition, we will assess statistical heterogeneity by examining the Higgins I^2^ statistic following these thresholds:

• < 25%: no (none) heterogeneity;
• 25% to 49%: low heterogeneity;
• 50% to 74%: moderate heterogeneity;
• ≥ 75%: high heterogeneity.

We will consider the following information for heterogeneity analysis:

#### Subgroup analysis

- Immune-mediated inflammatory disease (IMID) (e.g. Rheumatoid arthritis, Systemic Lupus, Spondyloarthritis, Sjögren syndrome,)
- Drug treatment (number of drugs, duration of drug treatment, dose, and type of drug treatment)
- Comorbidities
- Age
- Route of administration

#### Sensitivity analysis

- Studies with high risk of bias

### Assessment of reporting biases

When at least 10 studies are included in a meta-analysis we will explore the likelihood of reporting biases visually inspecting funnel plots. For continuous outcomes, Egger’s test will be used to detect possible small study bias as recommended in Cochrane Handbook for Systematic Reviews of Interventions.

## DISCUSSION

When the first reports about COVID-19 pathophysiology and clinical manifestations started to be published, there were some concerns with patients undergoing immunosuppressed treatment (3). However, unlike common viral agents, SARS-CoV-2 has not been shown to cause a more severe disease in patients with immune-mediated inflammatory diseases (5). To the best of our knowledge, the potential protective capacity of these drugs has not been properly evaluated and remains uncertain.

This rapid living review will systematically evaluate the best available evidence on the possible protective effect of drugs used in patients with immune-mediated inflammatory diseases for COVID-19. We expect it will help clinicians in their decision-making processes as, to date, no treatment has proven effective for treating COVID-19.

As we will follow the Cochrane Handbook of Systematic Reviews recommendations (7) and use extensive searches in the largest health databases, we believe we will be able to summarize the current available evidence and to clarify the potential of these drugs in preventing severe course of COVID-19.

## Data Availability

The authors confirm that the data supporting the findings of this study are available within the article

